# Association of Coronary Stenosis with Cerebral Small Vessel Diseases in Neurologically Asymptomatic Adults

**DOI:** 10.1101/2023.02.07.23285631

**Authors:** Kyungha Min, Jae-Moon Yun, Seo Eun Hwang, Ki-Woong Nam, Han-Yeong Jeong, Hyung-Min Kwon, Jin-Ho Park

## Abstract

**Background:** The high prevalence of coronary stenosis in patients with stroke is well established. However, the association between coronary stenosis and cerebral small vessel diseases (cSVD) in asymptomatic populations remains unclear.

**Methods:** As a cross-sectional study, we evaluated South Korean adults who underwent a health checkup including brain magnetic resonance imaging and coronary computed tomography angiography between January 2010 and December 2013. The degree of coronary stenosis was classified into three groups: no stenosis (0%), non-significant stenosis (1–49%), and significant stenosis (≥50%). cSVD includes silent lacunar infarction (SLI), cerebral microbleeds (CMB), and white matter hyperintensity (WMH). We used binary logistic regression analyses for SLI and CMB, and linear regression analysis for WMH.

**Results:** A total of 1,571 participants were evaluated. The prevalence of non-significant and significant coronary stenosis was 369 (23.5%) and 95 (6.1%), respectively. The prevalence of SLI and CMB was 112 (7.1%) and 66 (4.2%), respectively. The mean WMH volume was 2.6 ± 6.1 mL. SLI was significantly associated with both non-significant (adjusted odds ratio [aOR] = 1.94; *p* = 0.004) and significant coronary stenosis (aOR = 2.37; *p* = 0.011), even showing dose-response relationship (*p* for trend = 0.012). For WMH, only significant coronary stenosis was associated with WMH (β = 0.27; *p* = 0.013) and had a dose–response relationship (*p* for trend = 0.012).

**Conclusions:** The presence and severity of coronary stenosis were significantly associated with SLI and WMH. Physicians who detects any of coronary stenosis or cSVD should pay attention to the possible coexistence of the other disease.

## Introduction

Atherosclerotic cardiovascular disease (ASCVD) is a leading cause of death worldwide. According to the World Health Organization, an estimated 17.9 million people died from cardiovascular diseases (CVD), representing 32% of all global deaths in 2019. Of these, 85% were due to heart attack and stroke [1]. The burden of CVD in the number of disability-adjusted life years also continues to increase globally [2].

Stroke usually causes disabilities with limited neurological recovery; therefore, it is important to identify high-risk groups early, and manage modifiable risk factors [3]. Cerebral small vessel diseases (cSVD), which is a group of subclinical pathologies including silent lacunar infarction (SLI), cerebral microbleeds (CMB), and white matter hyperintensities (WMH), is known to lead to ischemic and hemorrhagic strokes [4]. Therefore, understanding the underlying mechanisms of cSVD development and its risk factors is essential for future stroke prevention.

The majority of patients with both nonfatal and sudden cardiac death do not experience any prior symptoms of chest pain or exertional dyspnea, highlighting the importance of early detection and treatment of underlying subclinical coronary atherosclerosis [5]. To date, growing evidence supports the use of noninvasive imaging tests to screen for coronary stenosis [6,7]. Coronary computed tomography angiography (CCTA) is an effective imaging technique that can identify plaque composition and the severity of coronary stenosis, and is increasingly accepted as a first-line test for the diagnosis of coronary artery disease [8]. Some studies have reported the effectiveness of CCTA screening for coronary artery disease in high-risk groups [9,10].

Several studies have shown a relationship between stroke and asymptomatic coronary stenosis, or cSVD and symptomatic coronary stenosis. The burden of asymptomatic coronary artery disease is high in patients with nonfatal cerebral infarction and unknown coronary heart disease [11]. Symptomatic coronary atherosclerosis is also a known risk factor for silent brain infarction [12]. Furthermore, significant associations between coronary artery calcification and cSVD in asymptomatic adults have also been reported [13]. Hence, we can assume that there may be a significant relationship between subclinical coronary stenosis and cSVD. However, to the best of our knowledge, no studies have investigated the relationship in neurologically and cardiologically asymptomatic screening populations. In South Korea, it is common to undergo health checkups using advanced imaging modalities such as brain magnetic resonance imaging (MRI) and CCTA. In this study, we aimed to evaluate the association between coronary stenosis and cSVD by using brain MRI and CCTA data in an asymptomatic screening population.

## Methods

### Study Population

We used a consecutive health checkup registry from the Seoul National University Hospital Health Promotion Center between January 2010 and December 2013. We retrospectively evaluated 1,627 subjects who underwent voluntary screening health checkups, including both brain MRI and concomitant CCTA. We excluded subjects who had reported a history of previous stroke, myocardial infarction, or angina (n=56). Finally, a total of 1,571 subjects who were neurologically and cardiologically asymptomatic were enrolled. This study was approved by the Institutional Review Board of the Seoul National University Hospital (IRB No. 1502-026-647). All experiments were performed in accordance with the Declaration of Helsinki and the relevant guidelines and regulations.

### Evaluation of Coronary Stenosis

CCTA was performed using a 64-slice dual-source CT scanner (Somatom Definition [Siemens Medical Solutions, Forchheim, Germany]). A computed tomography (CT) scan was performed using a retrospective ECG-gated mode with ECG pulsing. Scans were performed from the diaphragm to the level of tracheal bifurcation in the caudo-cranial direction. The means and ranges of the heart rates during CT scanning were recorded for all patients. The CT data was processed using a three-dimensional software (Rapidia [INFINITT, Seoul, Korea]), and the images were assessed by an experienced radiologist. The presence of coronary stenosis was evaluated semi-quantitatively and described as the greatest percentage of lumen diameter reduction among the three epicardial coronary arteries, including the left anterior descending artery, left circumflex artery, and right coronary artery (estimated as multiples of 5 %) [20,21]. If there was stenosis in two or more coronary arteries, the value with the highest degree of stenosis was selected. In the current study, we classified coronary stenosis into three groups: no stenosis (0%), non-significant stenosis (1-49%), and significant stenosis (≥ 50%).

### Evaluation of Cerebral Small Vessel Diseases

In this study, we evaluated all three components of cSVD: SLI, CMB, and WMH. The methods used to measure cSVD have been described in detail in previous studies [14]. In brief, brain MRIs with 1.5-or 3-Tesla MR scanners (Signa [GE Healthcare, Milwaukee, WI, USA] or Magnetom Sonata scanners [Siemens, Munich, Germany]) were performed in all subjects, and their brain MRI data were evaluated for cSVD. A SLI was defined as a focal infarction that was 3-15 mm in size and in the territory of the perforating branches to the basal ganglia, thalamus, internal capsule, corona radiata, centrum semiovale, brain stem, or cerebellum, which had a central signal intensity corresponding to the cerebrospinal fluid on T1- and T2-weighted images in MRI examinations [15]. A CMB was defined as a focal round lesion less than 10 mm in size with a low signal on T2-gradient echo images [16] and WMH volume was measured using a computer-assisted semi-automated technique (MIPAV version 7.3.0. (National Institutes of Health, Bethesda, MD). All radiological markers were rated by two neurologists and disagreements were resolved by discussion with a third rater.

### Clinical Variables

We measured demographic factors and clinical cerebrovascular and cardiovascular risk factors that were collected during the health checkup. Anthropometric factors, including height and weight, were measured while participants wore a light gown in an overnight fasting state. Body mass index (BMI) was calculated as weight (kg) divided by the square of height (m^2^) and the waist circumference (WC) was measured at the midpoint between the lowest rib margin and iliac crest in a standing position. Each participant’s lifestyle and known medical conditions were investigated through structured questionnaires and interviews by a trained family physician. Smoking status was classified as non-smoker, ex-smoker, or current smoker. Regular exercise was defined as moderate or high-intensity exercise for 150 min or more per week [17]. Information collected regarding medical status included previously diagnosed comorbid conditions, including stroke or myocardial infarction/angina, and use of related medications (i.e., antihypertensives, antidiabetic drugs, lipid-lowering drugs, anti-platelet drugs, and anti-coagulants). Blood pressure (BP) was measured after 5 min of rest in the sitting position. We also evaluated laboratory examination results including: fasting blood glucose, hemoglobin A1c (HbA1c), total cholesterol, triglyceride, high-density lipoprotein cholesterol, and low-density lipoprotein cholesterol. We calculated the estimated glomerular filtration rate (eGFR) using the Modification of Diet in Renal Disease (MDRD) formula. All serum samples were obtained after fasting for over 12 hours. Subjects were considered to have hypertension if they had a high systolic (≥140 mmHg) or diastolic (≥90 mmHg) blood pressure or were currently being treated with antihypertensive medications. Diabetes mellitus (DM) was defined as a fasting blood glucose level of 126 mg/dL or more or HbA1c more than or equal to 6.5%, or a self-reported prior diagnosis of DM with current use of insulin or oral hypoglycemic medications. Dyslipidemia was defined as a total serum cholesterol level of more than or equal to 240 mg/dL or current use of lipid-lowering drugs.

### Statistical analysis

The data is presented as means (SDs) or number of subjects (percentages), as appropriate. We transformed the WMH volume into a square root scale because of the excessive number of zeroes [18]. Descriptive statistics were used to assess basic characteristics of the study population. To conduct univariate and multivariate analysis, we used binary logistic regression analysis for SLI and CMB, and in the case of WMH, we conducted linear regression analysis. In the multivariate analysis, we included age, sex, BMI, smoking, exercise, hypertension, DM, and dyslipidemia as confounders because they are established cardiovascular risk factors [19]. According to previous studies, alcohol consumption was assumed to be a possible confounder [20]. Additionally, WC and eGFR were considered possible confounders because significant associations were observed in the univariate analyses. Therefore, we performed multivariate analyses adjusted for age, sex, BMI, WC, smoking, drinking, exercise, hypertension, DM, dyslipidemia, and eGFR. We also assessed the dose-response relationship between the degree of coronary stenosis and cSVD (*p* for trend). All statistical analyses in the current study were conducted using STATA version.16.1 (StataCorp, College Station, Texas, USA), and statistical significance was considered if the *p* value was less than 0.05.

## Results

We included a total of 1,571 neurologically and cardiologically asymptomatic participants (mean age, 56.7 years; female, 42.6%). Non-significant coronary stenosis was found in 369 (23.5%) participants, and significant coronary stenosis was observed in 95 (6.1%) participants. The prevalence of SLI and CMB were 112 (7.1%) and 66 (4.2%), respectively. The mean WMH volume was 2.6 ± 6.1 ml. With regards to smoking status, 828 (52.7%) participants were non-smokers, 446 (28.4%) were ex-smokers, and 297 (18.9%) were current smokers. There were 593 (37.8%) participants who were regarded as having hypertension, 259 (16.5%) participants had DM, and 358 (22.8%) had dyslipidemia. The other detailed baseline characteristics of the study subjects are presented in Table 1.

**Table 1.**
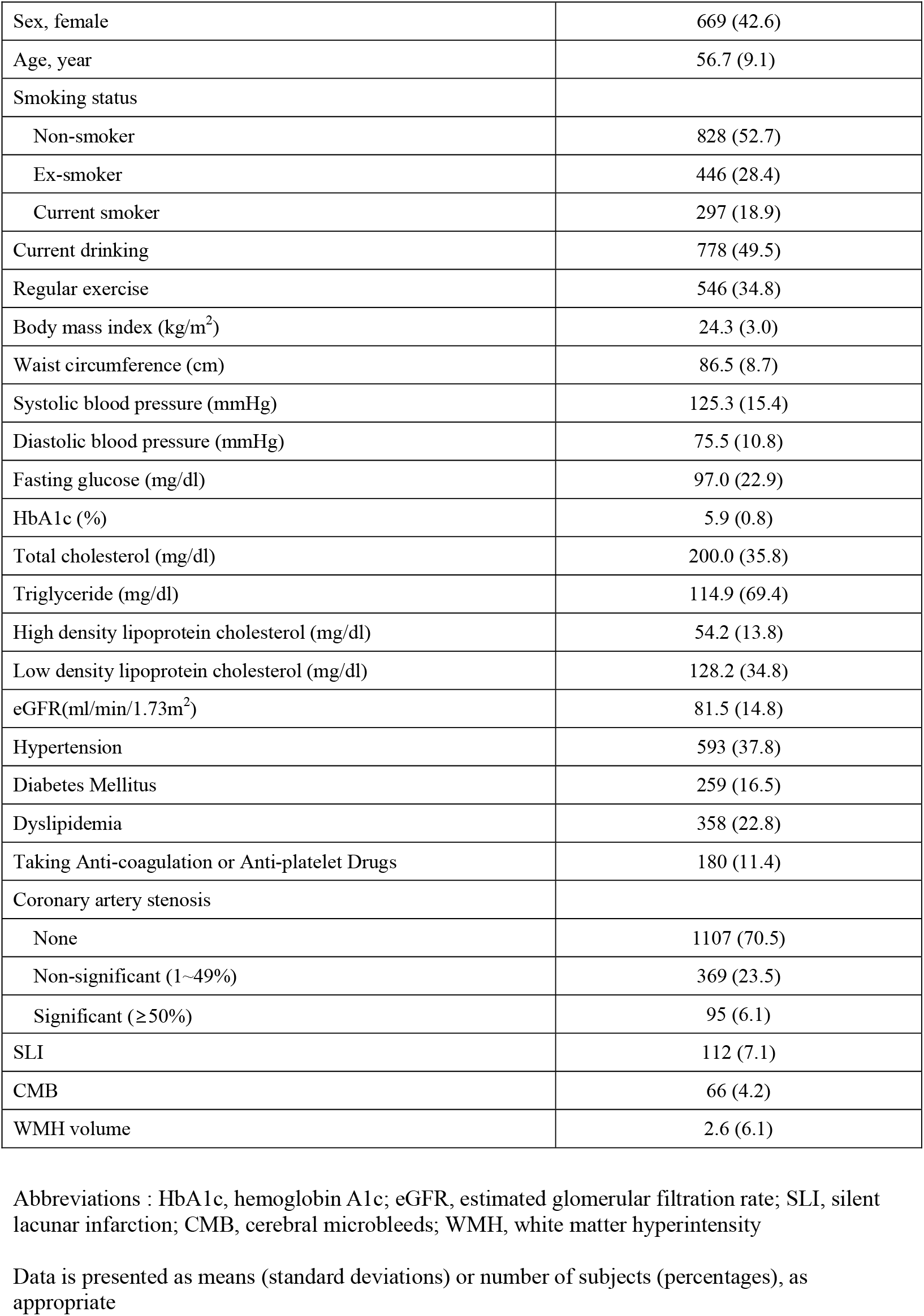
Baseline characteristics of subjects (n=1,571)

|  |  |
| --- | --- |
| Sex, female | 669 (42.6) |
| Age, year | 56.7 (9.1) |
| Smoking status |  |
| Non-smoker | 828 (52.7) |
| Ex-smoker | 446 (28.4) |
| Current smoker | 297 (18.9) |
| Current drinking | 778 (49.5) |
| Regular exercise | 546 (34.8) |
| Body mass index (kg/m<sup>2</sup>) | 24.3 (3.0) |
| Waist circumference (cm) | 86.5 (8.7) |
| Systolic blood pressure (mmHg) | 125.3 (15.4) |
| Diastolic blood pressure (mmHg) | 75.5 (10.8) |
| Fasting glucose (mg/dl) | 97.0 (22.9) |
| HbA1c (%) | 5.9 (0.8) |
| Total cholesterol (mg/dl) | 200.0 (35.8) |
| Triglyceride (mg/dl) | 114.9 (69.4) |
| High density lipoprotein cholesterol (mg/dl) | 54.2 (13.8) |
| Low density lipoprotein cholesterol (mg/dl) | 128.2 (34.8) |
| eGFR(ml/min/1.73m<sup>2</sup>) | 81.5 (14.8) |
| Hypertension | 593 (37.8) |
| Diabetes Mellitus | 259 (16.5) |
| Dyslipidemia | 358 (22.8) |
| Taking Anti-coagulation or Anti-platelet Drugs | 180 (11.4) |
| Coronary artery stenosis |  |
| None | 1107 (70.5) |
| Non-significant (1~49%) | 369 (23.5) |
| Significant (≥50%) | 95 (6.1) |
| SLI | 112 (7.1) |
| CMB | 66 (4.2) |
| WMH volume | 2.6 (6.1) |

Abbreviations : HbA1c, hemoglobin A1c; eGFR, estimated glomerular filtration rate; SLI, silent lacunar infarction; CMB, cerebral microbleeds; WMH, white matter hyperintensity
Data is presented as means (standard deviations) or number of subjects (percentages), as appropriate

In logistic regression analysis for SLI, non-significant coronary stenosis (adjusted odds ratio (aOR) = 1.94; 95% confidence interval (CI) = 1.23─3.05; *p* = 0.004) and significant coronary stenosis (aOR = 2.37; 95% CI = 1.22─4.63; *p* = 0.011) were found to be significantly associated with SLI after adjusting for possible confounders (Table 2). The severity of coronary stenosis was associated with an increasing trend in the prevalence of SLI (*p* for trend = 0.012).

**Table 2.**
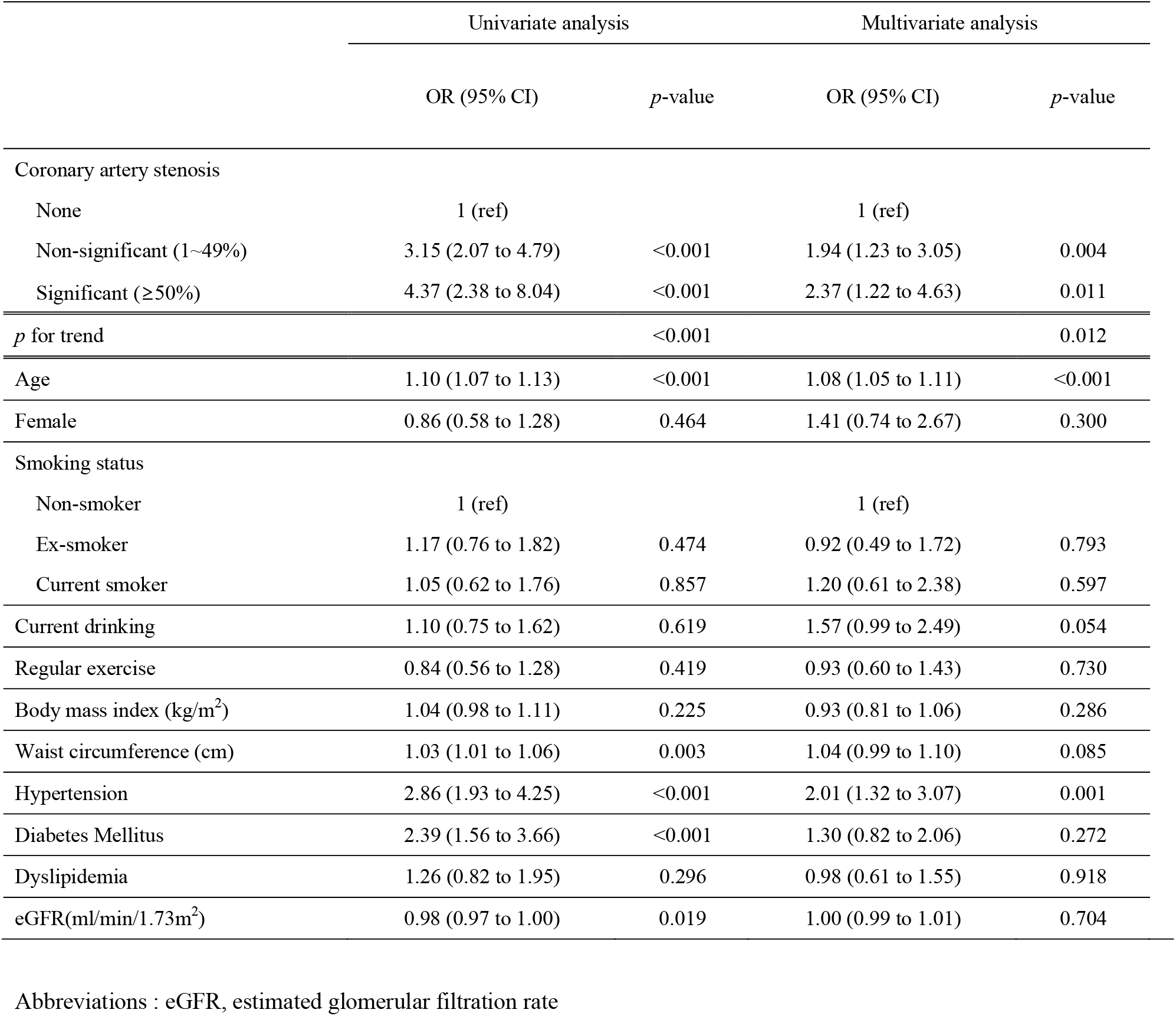
Simple and multiple logistic regression analysis between possible predictors and silent lacunar infarction

In the logistic regression analysis for CMB, although both non-significant (aOR = 1.43) and significant coronary stenosis (aOR = 1.18) were associated with a higher prevalence of CMB, statistical significance was not achieved (*p =* 0.216 and *p =* 0.728, respectively) (Table 3).

**Table 3.** Simple and multiple logistic regression analysis between possible predictors and cerebral microbleeds

|  | Univariate analysis |  | Multivariate analysis |  |
| --- | --- | --- | --- | --- |
|  | OR (95% CI) | *p*-value | OR (95% CI) | *p*-value |
| Coronary artery stenosis |  |  |  |  |
| None | 1 (ref) |  | 1 (ref) |  |
| Non-significant (1~49%) | 2.07 (1.22 to 3.52) | 0.007 | 1.43 (0.81 to 2.54) | 0.216 |
| Significant (≥50%) | 2.01 (0.82 to 4.89) | 0.126 | 1.18 (0.46 to 3.07) | 0.728 |
| *p* for trend |  | 0.126 |  | 0.703 |
| Age | 1.05 (1.02 to 1.08) | 0.001 | 1.04 (1.01 to 1.08) | 0.011 |
| Female | 0.66 (0.39 to 1.12) | 0.123 | 0.77 (0.35 to 1.69) | 0.512 |
| Smoking status |  |  |  |  |
| Non-smoker | 1 (ref) |  | 1 (ref) |  |
| Ex-smoker | 1.33 (0.76 to 2.33) | 0.312 | 0.87 (0.41 to 1.82) | 0.709 |
| Current smoker | 1.18 (0.61 to 2.28) | 0.630 | 0.96 (0.42 to 2.20) | 0.929 |
| Current drinking | 1.50 (0.91 to 2.47) | 0.114 | 1.68 (0.95 to 2.96) | 0.075 |
| Regular exercise | 0.87 (0.51 to 1.48) | 0.609 | 0.88 (0.52 to 1.52) | 0.656 |
| Body mass index (kg/m<sup>2</sup>) | 0.97 (0.89 to 1.05) | 0.405 | 0.97 (0.82 to 1.16) | 0.769 |
| Waist circumference (cm) | 1.00 (0.97 to 1.03) | 0.929 | 0.99 (0.93 to 1.05) | 0.685 |
| Hypertension | 2.48 (1.50 to 4.10) | <0.001 | 2.16 (1.27 to 3.65) | 0.004 |
| Diabetes Mellitus | 2.13 (1.23 to 3.70) | 0.007 | 1.61 (0.89 to 2.89) | 0.113 |
| Dyslipidemia | 0.74 (0.39 to 1.41) | 0.364 | 0.64 (0.33 to 1.23) | 0.184 |
| eGFR(ml/min/1.73m<sup>2</sup>) | 1.00 (0.98 to 1.01) | 0.635 | 1.00 (0.99 to 1.02) | 0.707 |

Abbreviations : eGFR, estimated glomerular filtration rate

According to linear regression for WMH, only significant coronary stenosis (*β* = 0.27; 95% CI = 0.06─0.48, *p =* 0.013) was found to be statistically significant. They also showed dose-response relationships (*p* for trend = 0.012) (**Table 4**).

**Table 4.** Simple and multiple linear regression analysis between possible predictors and white matter hyperintensity^*^

|  | Univariate analysis |  | Multivariate analysis |  |
| --- | --- | --- | --- | --- |
| | $\beta$ (95% CI) | *p*-value | $\beta$ (95% CI) | *p*-value |
| Coronary artery stenosis |  |  |  |  |
| None | 1 (ref) |  | 1 (ref) |  |
| Non-significant (1~49%) | 0.35 (0.22 to 0.47) | <0.001 | -0.02 (-0.14 to 0.11) | 0.800 |
| Significant ($\geq 50\%$) | 0.72 (0.49 to 0.94) | <0.001 | 0.27 (0.06 to 0.48) | 0.013 |
| *p* for trend |  | <0.001 |  | 0.012 |
| Age | 0.06 (0.05 to 0.06) | <0.001 | 0.05 (0.05 to 0.06) | <0.001 |
| Female | -0.09 (-0.20 to 0.02) | 0.094 | 0.02 (-0.13 to 0.16) | 0.847 |
| Smoking status |  |  |  |  |
| Non-smoker | 1 (ref) |  | 1 (ref) |  |
| Ex-smoker | 0.22 (0.09 to 0.34) | 0.001 | 0.15 (0.01 to 0.30) | 0.041 |
| Current smoker | -0.07 (-0.22 to 0.07) | 0.331 | 0.14 (-0.03 to 0.30) | 0.098 |
| Current drinking | -0.10 (-0.21 to 0.01) | 0.072 | 0.05 (-0.06 to 0.16) | 0.359 |
| Regular exercise | -0.12 (-0.24 to -0.01) | 0.038 | -0.11 (-0.21 to -0.01) | 0.040 |
| Body mass index (kg/m<sup>2</sup>) | -0.01 (-0.03 to 0.01) | 0.168 | -0.02 (-0.05 to 0.01) | 0.190 |
| Waist circumference (cm) | 0.00 (-0.00 to 0.01) | 0.203 | 0.00 (-0.01 to 0.01) | 0.748 |
| Hypertension | 0.43 (0.32 to 0.54) | <0.001 | 0.24 (0.14 to 0.35) | <0.001 |
| Diabetes Mellitus | 0.43 (0.29 to 0.58) | <0.001 | 0.14 (0.00 to 0.27) | 0.050 |
| Dyslipidemia | 0.01 (-0.12 to 0.14) | 0.846 | -0.13 (-0.24 to -0.01) | 0.035 |
| eGFR(ml/min/1.73m<sup>2</sup>) | -0.01 (-0.01 to -0.00) | <0.001 | -0.00 (-0.00 to 0.00) | 0.681 |

Abbreviations : eGFR, estimated glomerular filtration rate
\*These variables were transformed into square root scales.

## Discussion

This study aimed to investigate the association between coronary stenosis and cSVD in neurologically asymptomatic adults. There was a significant correlation with SLI and both non-significant and significant coronary stenosis, whereas, only significant stenosis was associated with WMH. Dose-response relationships were also observed for SLI and WMH. However, there was no relationship between coronary stenosis and CMB.

The significant association observed between coronary stenosis and SLI/WMH may have resulted from several mechanisms. First, since coronary stenosis and SLI/WMH share cardiovascular risk factors, such as hypertension, dyslipidemia, DM, and smoking, they may share the same pathophysiology to atherosclerosis. Comparing the results of the univariate and multivariate analyses in this study, it was found that the OR was attenuated when the risk factors were adjusted, indicating that they had shared risk factors. To date, the exact underlying etiology of cSVD is not well understood [4], but several explanations have been proposed. According to previous studies [21], cSVD may be affected by different pathologies. The most common form of small vessel pathology results from microatheroma at the origin of perforated arterioles, similar to large-vessel atherosclerosis. Failure of the blood-brain barrier, which causes leakage of serum components into and through the walls of cerebral small vessels, resulting in neuronal and glial damage, has also been suggested as an important cause of SVD-related pathology. According to a study [22], CMB is generally considered to have two types of vascular pathological changes: hypertensive vasculopathy and cerebral amyloid angiopathy. CMB is less associated with atherosclerosis, which may explain why there was no relationship between coronary stenosis and CMB in this study.

Second, endothelial dysfunction can cause a breakdown of the blood-brain barrier, resulting in perivascular tissue and arteriolar wall damage, which is considered important in coronary stenosis and SLI/WMH [4,23,24]. Third, oxidative stress resulting in chronic inflammation can lead to coronary stenosis and SLI/WMHs. Previous studies have shown that various noxious stimuli that induce vascular oxidative stress have been implicated in the pathogenesis of various disease states, including stroke [25] and coronary artery disease [26]. Fourth, environmental factors such as air pollution can lead to coronary stenosis and cSVD. One study suggested that increased exposure to air pollution is associated with cSVD [27] and coronary stenosis [28]. Additionally, genetic factors can also cause coronary stenosis [29] and cSVD [30].

In previous studies, researchers evaluated cSVD in patients with coronary artery diseases, such as myocardial infarction or angina. However, several studies have reported the prevalence of coronary artery disease in stroke patients. Uekita et al. showed that the prevalence of SLI was significantly greater in two- and three-vessel coronary artery disease groups than in zero- and one-vessel groups [31]. Hur et al. investigated 317 ischemic stroke patients with no known coronary heart disease and found that 48% of the patients had significant coronary stenosis [32]. However, until now, there has been a lack of research on the association between cSVD and coronary stenosis in neurologically and cardiologically asymptomatic populations. In addition to the above studies, our findings add strong evidence to the association between SLI/WMH and asymptomatic coronary stenosis. To the best of our knowledge, our study is the first to be conducted in a general screening population with a relatively large sample size. We also considered various risk factors, including lifestyle and disease factors, which may affect both cSVD and coronary stenosis.

The current study has several limitations. First, as a cross-sectional study, it could not prove causality between coronary stenosis and SLI/WMH. Therefore, careful interpretation is required. Second, because our study population was based on a health checkup registry, there may be potential for selection bias. Although our checkup center is one of the largest in South Korea and covers a nationwide population, our results need to be validated by population-based prospective cohort studies. Lastly, some subjects might have been taking medications that affect cSVD or coronary stenosis. In this study, we aimed to evaluate neurologically asymptomatic subjects and excluded subjects who experienced stroke or myocardial infarction/angina based on structured questionnaire results. Despite the exclusion criteria, there may be hidden effects from subjects taking antiplatelet and anticoagulant agents (i.e., aspirin, warfarin, clopidogrel, and other antithrombotic agents). If someone is neurologically asymptomatic, but takes one of these drugs for other reasons, such as atrial fibrillation or primary prevention for ASCVD, the underlying mechanism may be affected. Therefore, we conducted an additional analysis excluding 180 participants who were taking antithrombotic agents. This yielded the same results as the main analysis presented above **(Supp Table1)**.

Since coronary stenosis and SLI/WMH are closely related and share an underlying mechanism, people with SLI/WMH should be cautious about coronary artery disease and vice versa. Furthermore, individuals with SLI/WMH or coronary stenosis require appropriate management to prevent disease progression. It is necessary to manage risk factors and plan screening tests for high-risk groups for early detection. In particular, cSVD and coronary stenosis are treated by different specialists; therefore, they should be comprehensively managed.

In this study, we investigated the association between coronary stenosis and cSVD in neurologically and cardiologically asymptomatic adults. We found that both non-significant and significant coronary stenosis was associated with SLI and only significant stenosis was associated with WMH. Therefore, physicians who discover coronary stenosis or cSVD need to pay attention to the possible coexistence of the other disease, manage shared risk factors, and consider screening tests for the other diseases for early detection and management.

## Data Availability

All data for this paper could be supplied on reasonable request.

## Acknowledgements

None

## Sources of Funding

None

## Disclosures

This paper followed the Strengthening the Reporting of Observational Studies in Epidemiology (STROBE) statement [33].

## Tables

**Sup Table 1.** Multiple logistic/linear regression between possible predictors and silent lacunar

|  | SLI |  | CMB |  | WMH |  |
| --- | --- | --- | --- | --- | --- | --- |
| | OR (95% CI) | *p*-value | OR (95% CI) | *p*-value | $\beta$ (95% CI) | *p*-value |
| Coronary artery stenosis |  |  |  |  |  |  |
| None | 1 (ref) |  | 1 (ref) |  | 1 (ref) |  |
| Non-significant (1~49%) | 1.95 (1.18 to 3.24) | 0.010 | 1.78 (0.94 to 3.36) | 0.077 | 0.03 (-0.10 to 0.15) | 0.692 |
| Significant ($\geq 50\%$) | 2.24 (1.04 to 4.82) | 0.040 | 1.50 (0.52 to 4.32) | 0.453 | 0.26 (0.04 to 0.48) | 0.022 |
| Age | 1.09 (1.06 to 1.13) | <0.001 | 1.04 (1.00 to 1.08) | 0.047 | 0.05 (0.04 to 0.06) | <0.001 |
| Female | 1.19 (0.58 to 2.42) | 0.633 | 0.82 (0.34 to 1.95) | 0.651 | -0.01 (-0.16 to 0.15) | 0.924 |
| Smoking status |  |  |  |  |  |  |
| Non-smoker | 1 (ref) |  | 1 (ref) |  | 1 (ref) |  |
| Ex-smoker | 0.84 (0.42 to 1.67) | 0.620 | 0.71 (0.31 to 1.61) | 0.408 | 0.10 (-0.05 to 0.26) | 0.179 |
| Current smoker | 1.12 (0.52 to 2.38) | 0.778 | 1.06 (0.45 to 2.50) | 0.899 | 0.15 (-0.02 to 0.31) | 0.076 |
| Current drinking | 1.51 (0.91 to 2.51) | 0.112 | 1.97 (1.04 to 3.72) | 0.037 | 0.05 (-0.06 to 0.16) | 0.376 |
| Regular exercise | 0.93 (0.57 to 1.50) | 0.752 | 0.94 (0.52 to 1.70) | 0.835 | -0.14 (-0.25 to -0.04) | 0.008 |
| Body mass index (kg/m<sup>2</sup>) | 0.89 (0.76 to 1.04) | 0.155 | 0.87 (0.71 to 1.07) | 0.190 | -0.02 (-0.05 to 0.01) | 0.245 |
| Waist circumference (cm) | 1.05 (0.99 to 1.11) | 0.105 | 1.02 (0.95 to 1.09) | 0.579 | 0.00 (-0.01 to 0.01) | 0.958 |
| Hypertension | 2.29 (1.44 to 3.64) | <0.001 | 1.97 (1.11 to 3.52) | 0.021 | 0.23 (0.12 to 0.33) | <0.001 |
| Diabetes Mellitus | 1.27 (0.74 to 2.17) | 0.388 | 1.56 (0.79 to 3.08) | 0.202 | 0.17 (0.02 to 0.32) | 0.023 |
| Dyslipidemia | 1.30 (0.78 to 2.15) | 0.311 | 0.64 (0.30 to 1.34) | 0.233 | -0.13 (-0.25 to -0.01) | 0.031 |
| eGFR(ml/min/1.73m<sup>2</sup>) | 1.00 (0.99 to 1.01) | 0.867 | 1.00 (0.98 to 1.02) | 0.797 | 0.00 (-0.00 to 0.00) | 0.846 |

infarction/cerebral microbleeds/white matter hyperintensity\* excluding antithrombotic agents
Abbreviations : eGFR, estimated glomerular filtration rate; SLI, silent lacunar infarction; CMB, cerebral microbleeds; WMH, white matter hyperintensity
\*These variables were transformed into square root scales.

## Notes

### Competing Interest Statement

The authors have declared no competing interest.

### Funding Statement

Not applicable

### Author Declarations

This study was approved by the Institutional Review Board of the Seoul National University Hospital (IRB No. 1502-026-647).

